# Supporting Underrepresented Undergraduate Entry into Aging and Neurosciences Research and Clinical Careers: Student-rated Mentor Behaviors, Relationship Quality and Research Training Satisfaction

**DOI:** 10.64898/2026.04.15.26350982

**Authors:** Sheri Thompson, Lawrence Ong, Becky Marquez, Anthony J. A. Molina, Dennis R. Trinidad, Steven D. Edland, the MADURA Mentorship Program

**Affiliations:** Herbert Wertheim School of Public Health and Human Longevity Science, University of California San Diego, La Jolla, California, United States of America; Department of Neurosciences, University of California San Diego, La Jolla, California, United States of America; Department of Medicine, University of California San Diego La Jolla, CA, United States of America and Sam and Rose Stein Institute for Research on Aging, University of California San Diego, La Jolla, CA, United States of America

**Author notes:** Corresponding author (ST).

## Abstract

Improving diversity in U.S. Alzheimer’s disease (AD) research is a pressing need. By 2050, Hispanic and Latino Americans will comprise 30% of the population. Hispanics are 1.5 times more likely and Blacks are twice as likely to develop AD compared to Whites, yet both remain vastly underrepresented in clinical trials research. Aging and AD research mentorship of underrepresented STEM undergraduates is designed to promote entry into related professions by students committed to decreasing disparities in AD research participation and clinical care. The NIA-funded MADURA program recruited 93 students from backgrounds historically underrepresented in STEM majors and/or from NIH-defined disadvantaged backgrounds. Trainees were placed in aging/AD research labs and received weekly training and mentorship from faculty research PIs and other types of supervisors (postdoctoral researchers, graduate students, research assistant staff…) Our study examined student ratings of the program and mentor behaviors, using a program-specific survey and the Mentoring Competency Assessment-21 (MCA-21). Trainees were highly satisfied with both mentoring relationships and the overall program. Student rated MCA-21 competency areas were quite high for both P.I.s and other types of research mentors. However, there were striking differences in associations between competencies and relationship and program satisfaction, by mentor type. For PI mentors, no MCA-21 competencies were associated with relationship satisfaction, but five of six competencies were associated with relationship satisfaction for other mentor types. Similarly, no PI mentor competencies were significantly correlated with overall placement satisfaction, but all six competencies were correlated with overall placement satisfaction for other mentor types. The authors discuss the likelihood of differing student expectations of faculty PI versus other types of research mentors, recommendations for assessing role-specific student expectations (including functions primarily possible only for senior faculty PIs), and utilizing nearer-peer plus PI faculty mentors to comprehensively address the gamut of mentee needs.

## Introduction

### The need for diversity in aging and Alzheimer’s disease research

The demographics of the U.S. population are shifting dramatically, in terms of ethnic/cultural composition and age. As an example, the American Psychological Association notes that by 2050, Hispanic/Latino Americans (H/L) will comprise 30% of the U.S. population. [1] The H/L population age 65 and older was 4.6 million in 2019 and is projected to grow to 19.9 million by 2060. [2] Yet substantial barriers to health services and research opportunities continue, for this and many marginalized populations. A 2014 US Department of Health and Human Services literature review [3] reported “consistent and adverse disparities among Blacks and Hispanics compared to non-Hispanic whites in the prevalence and incidence of AD, mortality, participation in clinical trials, use of medications and other interventions, use of long-term services and supports, health care expenditures, quality of care and caregiving.” Although Hispanics are 1.5 times more likely and Blacks twice as likely to develop Alzheimer’s disease (AD), they and other minoritized groups are vastly underrepresented in AD clinical trials research. [4, 5]

The Social Determinants of Health (SDOH) was originally conceptualized in 1991, and research on individual health-related social needs suggests continued barriers which account for greater effects on health outcomes than clinical care per se and continue to drive health disparities. [6–8] These include economic constraints, limited education or health literacy, limited English proficiency, illness stigma, distrust of researchers and medical institutions, limited access to care, transportation issues, provider biases, and numerous other influences related to cultural differences and systemic inequities. Such underrepresentation is not an historical artifact. Research presented at the 2023 Alzheimer’s Association International Conference also notes new barriers for two recent clinical trials of monoclonal antibody AD medications. Specifically, Blacks and Hispanics wanting to participate in these drug studies were less likely to meet biomarker eligibility criteria due to lower levels of amyloid proteins at screening — and in the case of a donanemab trial, lower levels of tau proteins as well. Reardon notes that People of Color (a term she uses but does not formally define) comprised only 20% of participants in an international lecanemab trial and less than 10% in a donanemab trial. In the donanemab study specifically, Reardon reported that only 19 Black participants out of 1,736 total (barely more than 1%) participated in the trial. [9] Given myriad causes, underrepresentation in AD research studies perpetuates gaps in population-specific knowledge about the disease and treatment effects, which may vary by race, ethnicity and other characteristics.

### Lack of diversity in post-secondary and STEM education

Underrepresentation of post-secondary students of color has been documented for decades, in terms of both college degree attainment in general, and numerous specific areas of study. As an example, per the 2024 *State of U.S. Science and Engineering Report*, American Indian or Alaska Native, Black or African American, and Hispanic or Latino students were all underrepresented among science and engineering bachelor’s and graduate degree recipients. [10] Disparities have been demonstrated specifically in undergraduate and graduate neuroscience studies. Ramos and colleagues conducted an analysis using 30+ years of data from the National Center for Education Statistics, to assess demographic trends in neurosciences education participation. In 1980, there were fewer than 100 neuroscience graduates at any educational level. By 2015, there were more than 5000, 200, and 700 neuroscience graduates from undergraduate, master’s, and doctoral degree programs, respectively. Within the exponential growth from 1982 to 2012, underrepresented minority neuroscience graduates increased at every level. However, they comprised a vastly underrepresented proportion of the total neuroscience degree earners. Non-Hispanic Blacks earned only ∼4% and Hispanics earned only ∼8% of bachelor’s degrees between 1995 and 2015. Even as the total number of graduates at all levels increased, the rate of underrepresented minority student graduations remained relatively steady. [11]

### Mentorship to address educational and research disparities

At least for some SDOH and individual health-related social needs barriers, improved research inclusion may be most feasible if led by a highly trained, skilled workforce comprised - at least in proportionate numbers - by culturally aware researchers and allied care providers (impactful referral sources) who share some of the same lived experiences as the target populations. *Educational, research and career mentorship have gained attention as one means to enhance diversity in relevant STEM and medical professions, by building a more diverse workforce.* Many models have been implemented at levels ranging from pre-K through graduate studies and into career environments, with widespread support from federal institutions such as the National Institute of Medicine and the National Science Foundation [12] and the National Institutes of Health. [13] Research on post-secondary and graduate level research and STEM mentorship efforts, including our own program evaluations, have cited benefits including increased awareness of academic support resources, emotional support, improved professional self-efficacy, increased sense of belonging and instrumental networking in addition to skills development. [14–17] Quaye and colleagues assert that cultural and socio-emotional support systems are helpful for all students – but especially important for racial or ethnic minority students. Faculty mentors are likely to have greater impact for first-to-college students, who otherwise may lack plentiful role models for collegiate success. [18] In addition to gaining career-relevant education, undergraduate research and experiential learning have been identified as high impact practices for improving both URM STEM academic retention and success. [19]

### MADURA Program aims and structure

College mentorship program focus areas and target participants have varied widely, depending upon academic and community environments and funding sources. As a leading U.S. public university with strengths in research, neurosciences and health education, our UC San Diego team utilized funding from the National Institute on Aging to support diversity in aging and AD research and clinical careers. From 2019-2025, the MADURA (<u>M</u>entorship for <u>A</u>dvancing <u>U</u>ndergraduate <u>R</u>esearch on <u>A</u>ging) program served 93 undergraduates with these expressed career interests. All trainees met *either* racial or ethnic criteria for underrepresentation in STEM majors *and/or* met NIH Disadvantaged criteria. The primary Aim was to support these undergraduates in preparing for research or clinical careers related to aging or Alzheimer’s Disease.

After a competitive interview and matching process, undergraduate mentees completed 8 hours of paid weekly training and research work in faculty-led labs that aligned with student majors and career ambitions. They also participated in two weekly hours of paid whole-cohort research seminars and professional development trainings, as well as optional supplemental activities (attending conferences, developing and presenting research posters…) In contrast to Summer-focused training programs, MADURA mentees actively trained throughout the academic year and were encouraged to continue participation until attaining their baccalaureate degree. As such, a few mentees participated over multiple years.

The weekly MADURA Program curriculum was heavily informed by its ongoing trainee needs surveys, by quarterly surveys of participating mentors, by emerging literature on improving diversity in STEM postsecondary education and research, and resources from the National Academies of

Science, [14] the National Science Foundation [12] and the *Entering Research* training curriculum. [20] All of these sources also informed selection of four key mentor behaviors that were assessed through the program’s quarterly trainee surveys. With a strong commitment to process and outcomes evaluation, the MADURA Program collected anonymous quarterly and brief weekly student surveys plus quarterly surveys from research mentors that addressed both student-specific and general program performance. Survey data guided timely adaptations of group training content, such as identifying new topics sought by mentees or improved group training modalities. Recurring data collection assisted us in identifying top rated activities and presenters for continued inclusion. Likewise, trainees gave feedback on curriculum that merited improvement or lacked relevance to their current needs, allowing us to continually improve the training program. Surveys also provided key performance feedback – on student performance in research labs, and student perspectives on the performance of lab mentors. This paper focuses on the latter – examining mentor behaviors which were most beneficial to mentoring relationships and to trainees’ overall research training experience.

## Methods

### Student and Mentor Characteristics

Eligibility criteria for the MADURA mentorship program included being a matriculated undergraduate with a cumulative G.P.A. or 3.0 or higher, U.S. citizenship, reported interest in a career in aging or AD research or clinical services, ***AND*** belonging to one or more racial or ethnic groups underrepresented in science or engineering careers, relative to their proportion of the general population aged 20 to 34 years ***OR*** meeting two or more NIH Disadvantaged Criteria. Underrepresented groups from a 2024 NSF report (based upon 2021 data) 10 included: American Indian or Alaska Native, Black or African American, and Hispanic or Latino students. NIH Disadvantaged criteria are described as follows.

### Meeting at least two of the following criteria

1. Were or currently are homeless, as defined by the McKinney-Vento Homeless Assistance Act
2. Were or currently are in the foster care system, as defined by the Administration for Children and Families;
3. Were eligible for the Federal Free and Reduced Lunch Program for two or more years;
4. Have/had no parents or legal guardians who completed a bachelor’s degree (see the U.S. Department of Education);
5. Were or currently are eligible for Federal Pell grants;
6. Received support from the Special Supplemental Nutrition Program for Women, Infants and Children as a parent or child;
7. Grew up in one of the following areas: a) a U.S. rural area, as designated by the Health Resources and Services Administration Rural Health Grants Eligibility Analyzer, or b) a Centers for Medicare and Medicaid Services-designated Low-Income and Health Professional Shortage Areas (qualifying zip codes are included in the file).

*<u>Note</u>: Only one of the possibilities in #7 can be used as a criterion for the disadvantaged background.* [21]

Eligible applicants underwent competitive interviews for research lab openings appropriate for their major and specific career goals, if applicable. Top rated candidates were assigned to faculty-led research labs, until all current openings were filled. There were 17 participating research labs over the 2024-25 academic year. This year, a total of 24 student trainees participated in the MADURA program. Two students graduated mid-year and are therefore not included in Spring (end-of-year) analyses. Characteristics of the 2024-25 MADURA student cohort and the mentors’ research disciplines are described in **Tables 1 and 2**, below.

**Table 1.** 2024–25 Academic Year Mentee Cohort Characteristics (n = 24)

| Race | n | NIH Disadvantaged Criteria † | n |
| --- | --- | --- | --- |
| Hispanic/Latino | 12 | Not assessed † | 11 |
| Filipino | 5 | 2+ (meets NIH criteria) | 5 |
| Afghan | 1 | 1 or none | 8 |
| American Indian/Alaska Native | 1 |  |  |
| Black or African American | 1 |  |  |
| South Asian | 1 |  |  |
| Sri Lankan | 1 |  |  |
| Thai | 1 |  |  |
| Vietnamese | 1 |  |  |
| Academic Year | n | Academic Major | n |
| Senior | 15 | Human Biology | 7 |
| Junior | 8 | Neurobiology | 4 |
| Sophomore | 1 | Cognitive Behavioral Neuroscience | 4 |
|  |  | Public Health | 2 |
|  |  | Mathematics | 2 |
|  |  | Other Biology | 2 |
|  |  | Clinical Psychology | 1 |
|  |  | Social Psychology | 1 |
|  |  | Bioinformatics | 1 |
| Years of MADURA Participation | n |  | n |
| 1 to 1½ years | 15 |  |  |
| 2 years | 7 |  |  |
| 3 years | 2 |  |  |

† The Program did not originally utilize NIH Disadvantaged criteria, so it was not assessed for all applicants.

**Table 2.** 2024-25 Mentor/Supervisor Research Lab Disciplines (n=17)

| Discipline | Frequency * |
| --- | --- |
| Biology | 1 |
| Bioinformatics and/or Biostatistics | 2 |
| Medicine | 4 |
| Neurosciences | 8 |
| Pharmacology | 1 |
| Psychiatry | 4 |
| Public Health (Epidemiology) | 2 |

*\* Note: Three faculty conducted research projects combining more than one discipline.*

### Evaluation metrics

MADURA mentees completed quarterly digital evaluations of their experiences at research lab placements using a program-developed survey. This analysis utilized the most recent June 2025 cohort ratings. The **Quarterly MADURA Placement Experience Survey** prompted for a Likert scale satisfaction rating (1 = strongly disagree to 5 = strongly agree) with the mentoring relationship and the placement experience overall. Specific items were “I am generally satisfied with my Placement mentoring/supervisor relationship(s)” and “Overall, I am satisfied with my Placement activities.” In addition, mentees rated four specific mentor behaviors on a Likert rating scale of 1 = strongly disagree to 5 = strongly agree. <u>The assessed behaviors were</u>:

- Placement task assignments and expectations have been clearly explained.
- My mentor has provided honest feedback about my educational or career development plans.
- My mentor has provided constructive feedback about my performance at this placement.
- Assigned tasks and work opportunities included an appropriate balance of educational, training and/or challenging activities.

To deepen our understanding of the undergraduates’ research lab experiences, we also administered a validated STEM mentor behavior measure. The **Mentorship Competency Assessment-21 (MCA-21)** is a revalidation of an outcomes assessment tool developed for a prior multicenter RCT of the *Entering Mentoring* mentor training program. [22,23] The measure was initially designed to measure mentors’ pre- and post-training use of targeted *Entering Mentoring* program skills. The revalidated

MCA-21 is a 21-item scale which measures six mentor skill areas, with component alpha coefficients ranging from 0.77 to 0.86. Its developers assert that it effectively measures mentor competencies of **maintaining effective communication** (4 items), **aligning expectations** (4 items), **assessing understanding** (4 items), **fostering independence** (3 items), **addressing diversity** (3 items), **and promoting professional development** (4 items). [24] For both the original and revalidated MCA, one version may be completed by mentors as a form of *self*-evaluation. Another version allows *mentees* to rate their mentor on observed use of the same skills, allowing for comparison of mentor and mentee perceptions.

It should be noted that MADURA utilized the MCA-21 solely to gather information on mentor behaviors observed by students in their research labs, and *not* as an assessment of any program-provided mentor training. We also did not collect mentor self-ratings for comparison.

MADURA sought to learn which - if any - of the mentee-rated competencies were strongly associated with mentoring relationship and overall lab placement satisfaction. Many trainees received mentorship from both a lab’s Principal Investigator (PI) and another direct supervisor (typically a postdoc, graduate student or other research lab staff). Students were asked to complete MCA-21 ratings only for PIs and other supervisors with whom they averaged 4+ hours of contact time per week. This time could be comprised of any combination of 1:1 or group training, supervision or collaboratively completing research tasks. Per these criteria, some students rated only a PI mentor or only another type of direct supervisor, while some students rated both. Future references to “PIs” refers only to the faculty research lab Principal Investigators. “Direct Supervisors” refers to a variety of research lab personnel who were supplemental or sole providers of student mentorship, such as postdocs, research assistants and graduate students. For administration purposes, the MCA-21 items and Likert response scale were converted, verbatim, into a Qualtrics electronic survey administered in June of 2025.

At our research intensive institution, many trainees benefited from different types of mentors. A majority received mentorship from both a lab’s faculty Principal Investigator (PI) plus another direct supervisor (typically a postdoc, graduate student or other research lab staff). To ensure that trainees experienced adequate contact time to inform ratings, they were instructed to complete MCA-21 ratings *only* for PIs and Supervisors with whom they averaged 4+ hours of weekly contact time. This could be comprised of any combination of 1:1 or group training, supervision or collaboratively completing research tasks. Per these criteria, some students rated only a PI mentor or only another type of supervisor, while some students rated both. Future references to “PIs” refers only to the faculty research lab’s Principal Investigators. “Supervisors” refers to a variety of research lab personnel who were supplemental or sole providers of student mentorship, such as postdoc researchers, staff research assistants and graduate students.

### Analysis Plan

First, we used the quarterly MADURA Placement Experience Survey to evaluate potential associations between mentor behaviors and student satisfaction with the mentoring relationship and with their overall research placement experience. Next, we focused on the MCA-21 data. We calculated Cronbach’s Alpha to assess internal consistency of its six competency scales, for the PI and Supervisor ratings. This allowed us to investigate whether the MCA-21 performed similarly regardless of the type of mentor being rated, as we did not know if students might have differing expectations (and different internal rating criteria for specific behaviors) for faculty Principal Investigators versus other Supervisors.

Finally, we calculated Spearman correlation coefficients to examine potential associations of each MCA-21 competency area for PIs and Supervisors, to mentees’ mentoring relationship and research placement satisfaction. The FDR correction of 0.05 was used as a multiple testing correction to adjust the p-values for the Spearman correlation calculations. If a student left a question unanswered, the missing response was imputed with the mean of the other subscale items that assessed the same mentor competency. We conducted a sensitivity analysis to compare results with and without using imputed means, and obtained very similar Cronbach’s alpha coefficients, mean competency scores, correlation coefficients, and p-values. Therefore, the results of the analysis in this paper describe the competency subscale data with imputation of means used in downstream analyses.

## Results

Twenty-two students completed the June 2025 Quarterly MADURA Placement Experience Survey and the MCA-21 survey. For MCA-21 ratings, six students solely rated their PI, 8 students solely rated a supervisor, and 8 students rated both their PI and a Supervisor.

### Program-assessed Mentor Behaviors, Mentoring Relationship Satisfaction and Overall Placement Satisfaction

In terms of global satisfaction, student mentees rated both the mentoring relationship and overall placement satisfaction highly, with means of 4.727 and 4.636 respectively, on a Likert scale of 1 to 5. Relationship satisfaction was skewed extremely positively (SD=0.456, range of 4-5). Ratings of overall placement satisfaction were also skewed positively, with a standard deviation of 0.581 and range of 3-5. The most significant mentor behavior associated with both types of student satisfaction is “Assigned tasks and work opportunities included an appropriate balance of educational, training and/or challenging activities” with adjusted p-values of < 0.000 for both. Specific results follow in Table 3.

**Table 3.**
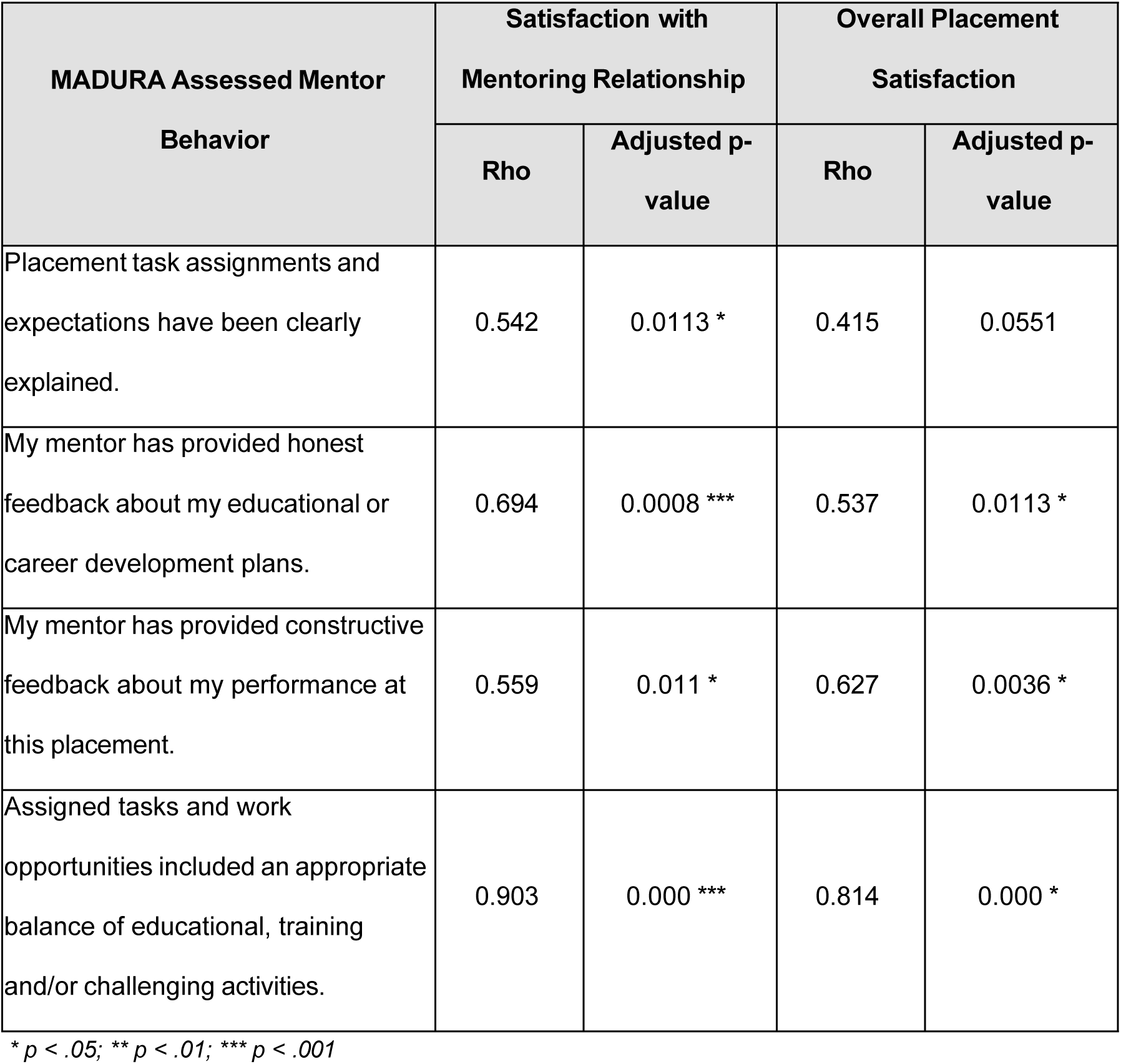
Spearman Correlations between MADURA Quarterly Survey Mentor Behaviors and Student Satisfaction Outcomes (June 2025; n=22)

### MCA-21 Internal Consistency

The MCA-21 survey assessed six mentor competencies through 3-4 questions per competency area. Cronbach’s Alpha scores were used to measure the effectiveness of items in a question set assessing the same mentor competency. As Table 4 illustrates, all PI mentor competencies had a Cronbach’s Alpha greater than .85, indicating good internal consistency. Internal consistency of the Supervisor ratings was even better, with the lowest Cronbach’s competency value > .85 and two above .95.

**Table 4.** Cronbach’s Alpha for Student Ratings of PI and Supervisor MCA-21 Competencies (June 2025)

| **Competency** | **Cronbach's Alpha for PIs**<br>**(n=14)** | **Cronbach's Alpha**<br>**for supervisors (n=16)** |
| --- | --- | --- |
| Effective Communication | 0.942 | 0.893 |
| Aligning Expectations | 0.917 | 0.956 |
| Assessing Understanding | 0.913 | 0.953 |
| Fostering Independence | 0.850 | 0.853 |
| Addressing Diversity | 0.898 | 0.870 |
| Promoting Professional Development | 0.912 | 0.924 |

### MCA-21 Competencies and Associations with Relationship and Overall Placement Satisfaction

As seen in Table 5, mentee ratings of Supervisor MCA-21 competencies were consistently quite high. For both PIs and Supervisors, all items had relatively small standard deviations and all values averaged 6.0 or higher (approaching the maximal value of 7 which was defined as “extremely skilled.”)

**Table 5.** Student Ratings of MCA-21 Mentor Competencies by Mentor Type (June 2025) Competencies related to Mentee Relationship Satisfaction and Research Placement Satisfaction.

| MCA-21 Mentor Competency | PI Mentor Ratings<br>(n = 14) |  | Supervisor Mentor<br>Ratings (n = 16) |  |
| --- | --- | --- | --- | --- |
|  | Mean | SD | Mean | SD |
| Effective Communication | 6.464 | 0.934 | 6.828 | 0.338 |
| Aligning Expectations | 6.143 | 1.099 | 6.640 | 0.626 |
| Assessing Understanding | 6.048 | 1.260 | 6.458 | 0.851 |
| Fostering Independence | 6.310 | 0.947 | 6.563 | 0.805 |
| Addressing Diversity | 6.369 | 0.934 | 6.667 | 0.656 |
| Promoting Professional Development | 6.339 | 0.973 | 6.328 | 1.182 |

As illustrated in Table 6, there were striking differences in the associations between MCA competencies and relationship and placement satisfaction, for PIs and supervisors. None of the MCA-21 assessed competency areas were associated with relationship satisfaction for PIs, while all but one competency area was associated with relationship satisfaction for Supervisors. As seen in Table 7, none of the PI competencies were correlated with overall research placement satisfaction, while all six supervisor competencies were significantly associated with research placement satisfaction.

**Table 6.** Spearman Correlations between MCA-21 Competencies and Mentoring Relationship Satisfaction by Mentor Type (June 2025)

| MCA-21 Competency | PI Relationship Satisfaction (n = 14) |  | Supervisor Relationship Satisfaction (n = 16) |  |
| --- | --- | --- | --- | --- |
|  | Rho | Adjusted p-value | Rho | Adjusted p-value |
| Effective Communication | 0.472 | 0.088 † | 0.596 | 0.020 * |
| Aligning Expectations | 0.369 | 0.194 † | 0.480 | 0.060 † |
| Assessing Understanding | 0.487 | 0.077 † | 0.707 | 0.004 ** |
| Fostering Independence | 0.460 | 0.098 † | 0.846 | 0.000 *** |
| Addressing Diversity | 0.379 | 0.181 † | 0.557 | 0.027 * |
| Promoting Professional Development | 0.477 | 0.084 † | 0.707 | 0.004 ** |

\* $p < .05$ ; \*\* $p < .01$ ; \*\*\* $p < .001$ ; † N.S.

**Table 7.** Spearman Correlations between MCA-21 Competencies and Overall Placement Satisfaction by Mentor Type (June 2025)

| MCA-21 Competency | PI Mentor Correlations (n = 16) |  | Supervisor Mentor Correlations (n = 16) |  |
| --- | --- | --- | --- | --- |
|  | Rho | Adjusted p-value | Rho | Adjusted p-value |
| Effective Communication | 0.472 | 0.088 † | 0.653 | 0.009 ** |
| Aligning Expectations | 0.369 | 0.194 † | 0.566 | 0.027 * |
| Assessing Understanding | 0.487 | 0.077 † | 0.806 | 0.001 ** |
| Fostering Independence | 0.460 | 0.098 † | 0.741 | 0.003 ** |
| Addressing Diversity | 0.379 | 0.181 † | 0.652 | 0.009 ** |
| Promoting Professional Development | 0.477 | 0.084 † | 0.788 | 0.001 ** |

\* $p < .05$ ; \*\* $p < .01$ ; \*\*\* $p < .001$ ; † N.S.

## Discussion

### Quarterly MADURA Survey of Mentor Behaviors

As assessed by the Quarterly Program survey, mentor behaviors of clear task explanations, honest educational and career feedback, constructive performance feedback and providing balanced activities for trainees were all positively associated with mentoring relationship satisfaction. We had originally targeted these behaviors for assessment, due to their reported importance in URM mentoring literature. Their strong associations with mentee relationship satisfaction support their utility as mentoring relationship quality indicators. Further, three of the four behaviors (honest educational and career feedback, constructive performance feedback and providing balanced activities for trainees) were also associated with overall research placement satisfaction. These are therefore appropriate mentor training targets, with balance in assigned activities (a mix of educational, skills training and novel or challenging tasks) most strongly associated with students’ overall research experience satisfaction. Although it trended towards significance, receiving clear instructions was not significantly associated with overall placement satisfaction. Although clear task explanations are a pre-requisite to enabling trainees to perform good work, the other three assessed behaviors are more central to the students’ perception of overall lab experience quality. The results suggest that our trainees perceived the need to request additional task explanations as normative and/or not substantially influential on perceptions of overall research experience quality. Based upon our findings, we suggest that PIs and supervisors carefully attend to providing comprehensive and increasingly more demanding experiences that expand student skill sets, to support both optimal mentoring relationships and research experience quality. Adequate skills training and foundational education are also vital, but trainee satisfaction also hinges upon opportunities to sequentially build skills and knowledge.

### Assessing MCA-21 Competencies

The high internal consistency of all six Competency scales was impressive and added to the evidence that the selected items measure their targeted classes of mentor behaviors. Regarding the associations between MCA-21 Competency Areas and Mentoring Relationship and Overall Placement Satisfaction: We were surprised to see that none of the PI MCA-21 Competency areas were significantly correlated with students’ mentoring relationship satisfaction, nor the overall research placement experience. In contrast, for other types of mentors, five of six MCA-21 competencies were significantly associated with mentoring relationship satisfaction and all six were correlated with overall placement satisfaction. As previously mentioned, direct supervisors were typically postdoc researchers, graduate students, research assistants or other research staff. Supervisors were highly competent mentors, possessing substantially greater research expertise than the undergraduate mentees. However, compared to well-established faculty research PIs, these other supervisors were perceived as more similar in educational level, research skills and/or authority. As such, these supervisors were often reported by mentees to be highly approachable, accepting, empathic and cognizant of trainee’s current knowledge and abilities. In labs that offered support from both PIs and other research team members, it was other types of supervisors that mentees reporting accessing first with additional training needs or instruction.

Prior verbal and assessment feedback on student-reported benefits of MADURA suggests that they tend to seek different benefits from connections with PIs. Specifically, our trainees have looked to PIs for critical support functions that require PI-level expertise, networks and authority. Examples include providing recommendations for jobs and graduate programs, activating deep professional networks to find employment opportunities for students, making compelling referrals for internships and other training programs, financially supporting conference attendance, and extending invitations to co-author manuscripts and presentations. [17] Per five years of quarterly MADURA surveys, mentees looked to PI faculty for these impactful functions and frequently cited them as their top-rated program benefits. Two kinds of support that might be more practical for PI-level faculty mentors are represented by MCA-21 items about helping mentees to network and acquire resources. [24] However, most of the other PI related professional development support functions most highly rated by MADURA trainees are not assessed by the MCA-21 professional development competency area. [17]

Our results suggest that the MCA-21 captures many of the key competencies needed by direct supervisors for optimal relationships and overall satisfaction. In contrast, mentees in multi-level research settings may evaluate relationship and placement satisfaction using a variety of role-specific criteria not represented by the MCA-21, especially as related to satisfaction with the faculty PI mentoring relationship and research experience. Students at different educational junctures may also expect different types of support from PIs. Drawing examples from five years of MADURA training, a senior applying to graduate school might have considered PI facilitation of scientific conference attendance as a major determinant of relationship and placement satisfaction. A new transfer student may prioritize educational and career feedback as the top need from a PI, while a graduating senior prioritizes receiving faculty recommendation letters or job referrals.

To summarize, student mentee expectations appear to vary by mentor type. It appeared that our mentees expected supervisors to utilize the full scope of competencies measured by the MCA-21, while looking to PI mentors for a narrower range of very important, stage-specific functions. Further research should evaluate potential differences in students’ definitions of optimal mentoring from research lab PIs, versus immediate supervisors. Mentorship programs with adequate bandwidth should conduct baseline surveys of mentee expectations of different types of research mentors. This information could assist program administrators to align expectations, and program developers and mentors to better address heterogeneous trainee needs. Because they are valued PI capabilities, active support with networking, referrals and tangible research support (funding conference participation, extending co-authorship opportunities…) should be among possible trainee needs to be assessed.

### Limitations

These analyses of associations between quarterly survey mentor behaviors and relationship and placement satisfaction, and of MCA-21 competencies and relationship and placement satisfaction – were correlational and cannot prove causation. Also, the student mentee surveys were anonymous, to alleviate student concerns over feedback reaching mentors. However, it is possible that some mentees may have been overly favorable in their ratings, due to fear of being identified and/or gratitude for funding, research training, and career support. Students were presumably aware that anonymous electronic survey administration and the size of our cohort would have make it difficult or impossible to identify individual respondents, so these survey results likely accurately reflect most trainees’ perceptions. Still, possible mentee biases cannot be definitely ruled out.

### Suggested Mentorship Program components

Based upon MADURA quarterly surveys, the most powerful factor associated with mentee relationship and placement satisfaction was a balance of skills training, research education and opportunities to expand skills with challenging and novel tasks. Research-focused mentorship programs should take special care to ensure that all three components are provided. Additionally, providing honest feedback about mentee educational and career development plans, and constructive feedback about student performance significantly impacts student placement satisfaction. Programs should incorporate regular feedback mechanisms on these topics, such as recurring semi-structured student evaluations, independent development plan (IDP) collaboration and routine updates, and 1:1 and group supervision.

Our findings also reinforce the tremendous value of “nearer” peer mentorship to undergraduate mentees’ research experiences. The benefits of cognitive and social congruence (perceived similarities) for enhanced learning have been described by other researchers. [25, 26] In the case of MADURA, supervisors were highly competent and more advanced in their research and educational careers than undergraduate mentees. Yet these research assistants, graduate students and/or postdocs were typically closer in educational status, experience, authority and/or career stage than well-established faculty Principal Investigators. Trainees reported greater comfort disclosing questions, errors and deficits, to these supervisors. These supervisors were especially well suited to empathize and collaborate on managing current barriers and pressures impacting mentee performance. For research labs with multi-level teams, combining mentorship from both the highly esteemed and well-connected faculty and nearer-peer supervisors confers a wider range of benefits than mentorship from either alone. We encourage research PIs to ensure involvement of graduate and other supervisors/trainers in undergraduate mentorship, pairing undergrad mentees with nearer-peers and engaging them in multi-level team projects and meetings, whenever possible.

## Data Availability

De-identified quantitative data from participant surveys has been made accessible via Zenodo at https://zenodo.org/doi/10.5281/zenodo.19489880.

